# Exome-wide association studies discover germline mutation patterns and identify high-risk populations in human cancers

**DOI:** 10.1101/2022.06.11.22275897

**Authors:** Sipeng Shen, Yunke Jiang, Guanrong Wang, Hongru Li, Dongfang You, Weiwei Duan, Ruyang Zhang, Yongyue Wei, Hongbing Shen, Zhibin Hu, David C. Christiani, Yang Zhao, Feng Chen

## Abstract

Genome-wide association studies have discovered numerous common variants associated with human cancers. However, the contribution of exome-wide rare variants to cancers remains largely unexplored, especially for the protein-coding variants. The UK Biobank provides detailed cancer follow-up information linked to whole-exome sequencing (WES) for approximately 450,000 participants, offering an unprecedented opportunity to evaluate the effect of exome variation on pan-cancer. Here, we performed exome-wide association studies (ExWAS) based on single variant levels and gene levels to detect their associations across 20 primary cancer types in the discovery set (WES-300k, N = 284,456) and replication set (WES-150k, N = 143,478), separately. The ExWAS detected 143 independent variants at variant-level and 49 genes at gene-level, while nine variants and eight genes were shared across cancers. In the cross-trait meta-analysis, we identified 239 additional independent pleiotropic variants, mapping to the genes which were functional through trans-omics analyses in transcriptomics and proteomics. Further, we developed exome-wide risk scores (ERS) to identify high-risk populations based on rare variants with minor allele frequency (MAF) < 0.05. The ERS had satisfactory performance in cancer risk stratification, especially for the extremely high-risk persons (top 5% ERS) that were frequently risk allele carriers. The ERS (median C-index (IQR): 0.655 (0.636-0.667)) outperforms the traditional polygenic risk score (PRS) (median C-index (IQR): 0.585 (0.572-0.614)) for discrimination in the replication set. Our findings offer further insight into the genetic architecture of human exomes for cancer susceptibility.

## Introduction

Cancer ranks as a leading cause of death and a critical barrier to increasing life expectancy worldwide ^1^. Population-based early screening approaches showed a remarkable reduction in cancer mortality ^2^, such as low-dose computed tomography (CT) screening for lung cancer ^3,4^. Considering the cost-effectiveness balance, it is generally agreed that screening should be limited to the high-risk population. However, precisely identifying high-risk persons is still challenging, while cancer is a complex disease that derives from environmental exposure and inherent heredity ^5^.

Cancer shows substantial heritability from genetic variants ^6^. Genome-wide association studies (GWAS) have identified numerous associations of genome-wide significance between genetic variants and common diseases ^7^. However, common single nucleotide polymorphisms (SNPs) identified in GWAS explain only a small fraction of heritability, which might be limited to the coverage of SNP arrays ^8^. Exome-wide association studies (ExWAS) have shown that rare coding variants tend to have larger phenotypic effects than common SNPs and contribute an essential component of heritability ^9^. Due to the effect allele frequency being generally low in ExWAS, the sample size should be large (e.g., *n* > 100,000) to guarantee the statistical power, especially for the rare variants ^10,11^.

Moreover, it is widely recognized that the polygenic risk score (PRS) is a powerful tool to discriminate the high-risk population susceptible to specific cancer ^12,13^. However, most PRSs are generated using common variants derived from the SNP array, which ignore the rare variants with larger effects, especially those located on exomes with remarkable biological significance ^14,15^. Thus, the rare variants might provide complementarity value for cancer risk stratification based on traditional PRS.

The UK Biobank (UKB) is a powerful resource for evaluating the associations between coding variants and human diseases because of its large sample size with high-quality whole-exome sequencing (WES) data (*n*≈450,000) ^16,17^. In our study, we investigated the landscape of genetic variants with multiple primary cancers through UKB WES project. Further, we leveraged the rare variants to improve the risk stratification models to identify the high-risk population.

## Results

### Exome-wide association study for single variants

Our study included the whole-exome sequencing 450k (WES-450k in data-field 23148) population of European ancestry in UK Biobank (Table S1). To ensure the robustness of the results, we conducted a two-stage association study in two separate datasets: discovery set (interim WES-300k in data-field 23146, N = 284,456) and replication set (the remaining WES-150k, N = 143,478). Overall, our study included 20 cancer types containing 106,836 primary cancer cases and 321,098 shared cancer-free controls. The number of cancer cases ranged from 773 (thyroid cancer) to 32,307 (skin cancer). 1,769,329 exome single nucleotide variants (SNVs) annotated putative loss-of-function (LoF), missense, and synonymous passed the quality control procedures.

For single-variant analyses, 306,031 variants with minor allele count (MAC)≥10 were tested in ExWAS. The genomic inflation factor (GIF) values suggested no obvious population stratification (Figure S1). When combining all the *P* values of pan-cancer, the GIF was 1.105, indicating the existing pleiotropic effects (Figure S2).

Among the 20 cancer types, ExWAS detected 255 signals from 242 variants that passed the genome-wide significance level (*P* < 5×10^−8^) in 12 cancer types. After LD pruning, we observed 153 independent signals across 64 chromosome regions (LD-*r*^*2*^<0.5) (Figure 1a, Table S2). The top variant with maximum association signals was rs555607708 (22:28695868:AG:A, frameshift variant of *CHEK2*, HGVSp: p.Thr367fs, effect allele frequency: 0.22%), which was associated with three cancer types including breast [OR (95% CI): 4.15 (3.16-5.43), *P* = 7.24×10^−25^], prostate [OR (95% CI): 2.40 (1.82-3.16), *P* = 3.48×10^−10^] and leukemia [OR (95% CI): 8.48 (4.12-17.48), *P* = 1.21×10^−9^].

**Figure 1.**
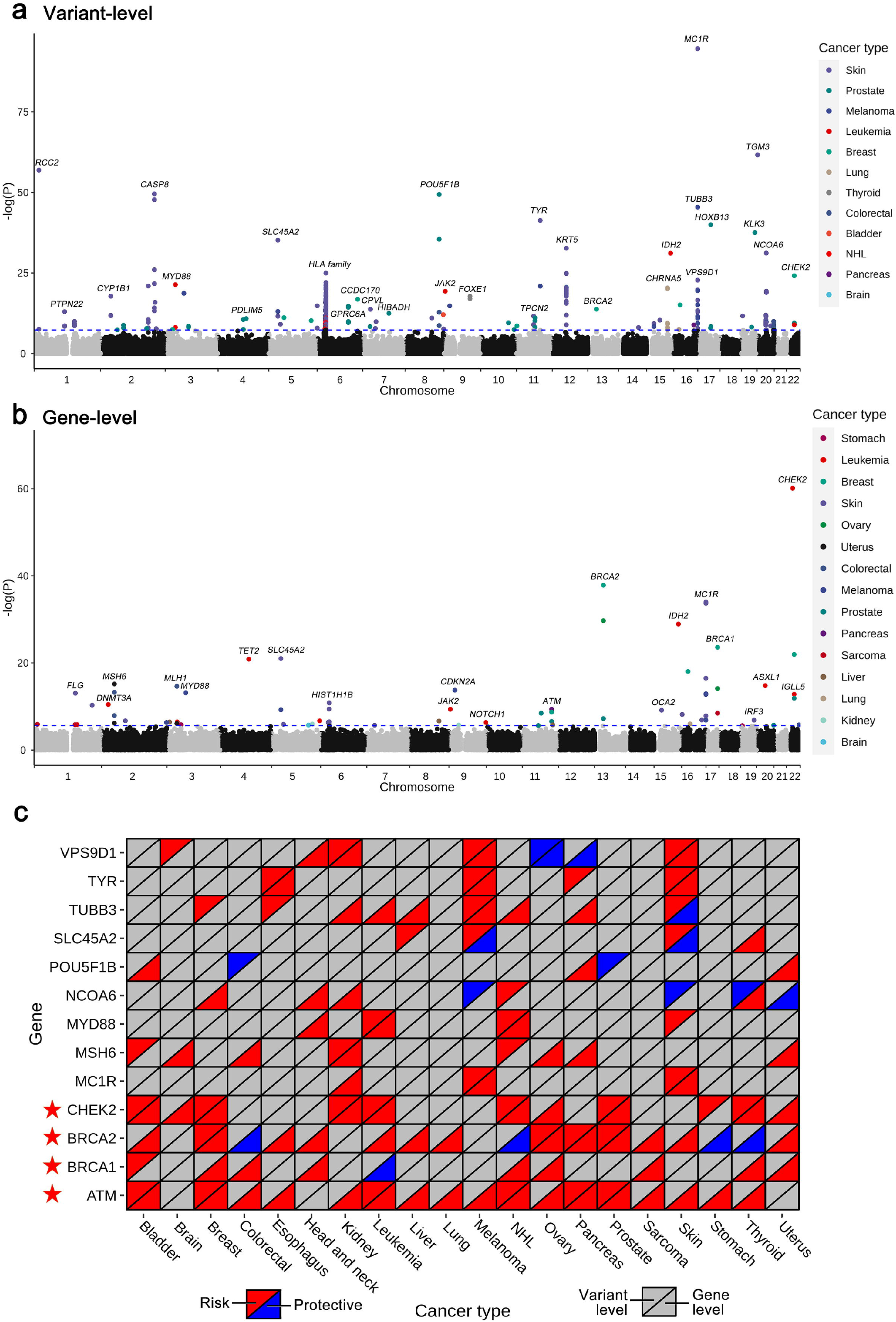
(a) Manhattan plot for the single variant association results in ExWAS. The blue dash line indicates the genome-wide significance level (*P* < 5× 10^−8^). (b) Manhattan plot for the gene-based association results in ExWAS. The blue dash line indicates the Bonferroni correction level (*P* < 2.5 × 10^−6^). (c) The heatmap of variant-level and gene-level association results for the genes shared at least two cancer types. We report the signal if it reaches nominal *P*<0.05 in the corresponding cancer type (red: odds ratio (OR)>1; blue: OR<1). The grey color indicates association *P* > 0.05.

Additionally, eight independent variants had significant associations with at least two cancer types, leading by rs6998061 (OR=0.82∼0.89, missense variant in *POU5F1B*), rs16891982 (OR=1.42∼1.46, missense variant in *SLC45A2*), rs387907272 (beta=24.1∼76.2, LoF variant in *MYD88*), rs3787220 (OR=0.85∼0.88, synonymous variant in *NCOA6*), rs77681059 (OR=1.11∼1.32, missense variant in *TUBB3*), rs1805007 (OR=1.34∼1.64, missense variant in *MC1R*), rs56288641 (OR=1.28∼1.59, missense variant in *VPS9D1*), rs1126809 (OR=1.13∼1.23, missense variant in *TYR*). More than half of the variants were shared between skin cancer and melanoma, including three variants located on 16q24.3. The remaining three variants were shared among breast, prostate, leukemia, colorectal cancers, and non-Hodgkin’s lymphoma (NHL) (Table S3).

### Exome-wide association study based on gene-level

In addition to the variant-level analyses, we performed gene-based association studies to capture the effects of ultra-rare variants. We used different genetic models to detect the signals, according to minor allele frequency threshold (0.05, 0.01, 0.001) and variant functional annotation (LoF, LoF+missense, LoF+missense+synonymous). After Bonferroni correction, 49 genes were considered significant (*P* < 2.5×10^−6^) (Figure 1b, Table S4). The top genes with the highest hit frequency were *CHEK2* [breast (OR = 1.03, *P* = 1.08×10^−22^), prostate (OR = 1.02, *P* = 1.28×10^−12^), leukemia (OR = 1.04, *P* = 1.55×10^−13^)], *BRCA2* [breast (OR = 1.08, *P* = 1.43×10^−38^), ovary (OR = 1.14, *P* = 1.94×10^−30^), prostate (OR = 1.04, *P* = 5.98×10^−8^)], and *ATM* [breast (OR = 1.10, *P* = 4.33×10^−10^), prostate (OR = 1.04, *P* = 1.89×10^−9^), pancreas (OR = 1.04, *P* = 2.67×10^−7^)], which were associated with three cancer types. *VPS9D1, SLC45A2, BRCA1, MSH6*, and *MC1R* were associated with two cancer types (Table S5).

We summarized the association results of genes that reached significance level in at least two cancer types from variant-level and gene-level analyses (Figure 1c). Four genes, including *CHEK2, ATM*, and *BRCA1/2*, showed a close relationship with human cancers.

### Shared genetic cross-trait meta-analyses identify pleiotropic signals

To identify additional potential pleiotropic variants associated with multiple cancers, we performed a cross-trait meta-analysis using Association analysis based on SubSETs (ASSET). 1,572 variants that reached *P* < 10^−4^ in any cancer type were included. We identified 150 independent variants (mapping to 86 genes, 43 chromosome cytobands) with significant one-directional effects (Table S6) and 89 independent variants (mapping to 38 genes, 16 cytobands) with significant bidirectional effects (Table S7) (Figure 2a).

**Figure 2.**
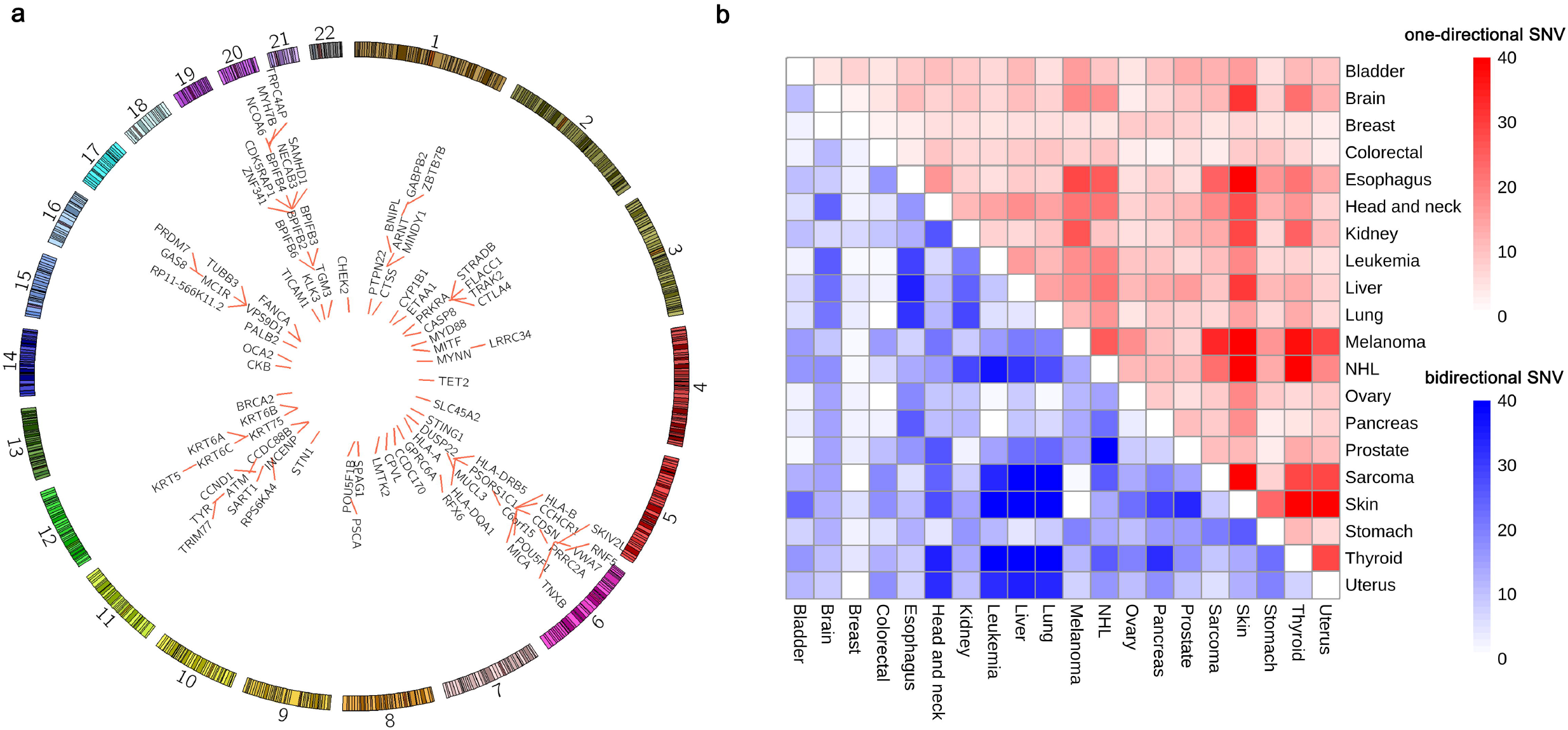
(a) Circos plot for the genes identified in the cross-trait meta-analysis. (b) Heatmap for the number of shared genetic variants across each cancer pair. The red color indicates the shared variants with one-directional effects. The blue color indicates the shared variants with bidirectional effects between the cancer pairs.

We checked the pleiotropic variants in NHGRI-EBI GWAS Catalog ^18^, a publicly available database that collected all published GWAS signals phenome-wide. The due date of associations collection was April 7, 2022. More than half of the identified variants were not reported in GWAS Catalog [100 novel one-directional variants (66.7% of all), 59 novel bidirectional variants (66.3% of all)] (Table S6-S7).

We also summarized the shared genetic variants across cancers identified in ASSET subgroups according to their effect direction (Figure 2b). The top cancer pairs with the highest shared one-directional variants were skin & melanoma (79 variants), skin & thyroid (69 variants), sarcoma & skin (51 variants), NHL & skin (50 variants), esophagus & skin (47 variants). The top cancer pairs with the highest heterogeneous bidirectional variants were leukemia & thyroid (51 variants), liver & thyroid (50 variants), lung & sarcoma (48 variants), skin & liver (48 variants), lung & thyroid (48 variants) (Table S8).

### Trans-omics functional analysis for the identified genes

We performed a trans-omics analysis to integrate the identified genes from single variant, gene based and cross-trait meta-analyses into transcriptomics and proteomics. Gene expression was obtained from the recompute transcriptomic data of The Cancer Genome Atlas (TCGA) and The Genotype-Tissue Expression (GTEx). 98 unique protein-coding genes across 15 tissue types that passed quality control were included. Through comparisons of gene expression in tumor and healthy normal tissues, we observed remarkable differences in gene expression. The well-known *HLA* family genes, *BRCA1/2, CHEK2*, and *TUBB3* were up-regulated in tumor tissues, while *ATM, VPS9D1*, and *MC1R* were down-regulated. However, some genes showed heterogenous trends across cancers, such as the *KRT* family, *POU5F1B*, and *TET2* (Figure 3a, 3b).

**Figure 3.**
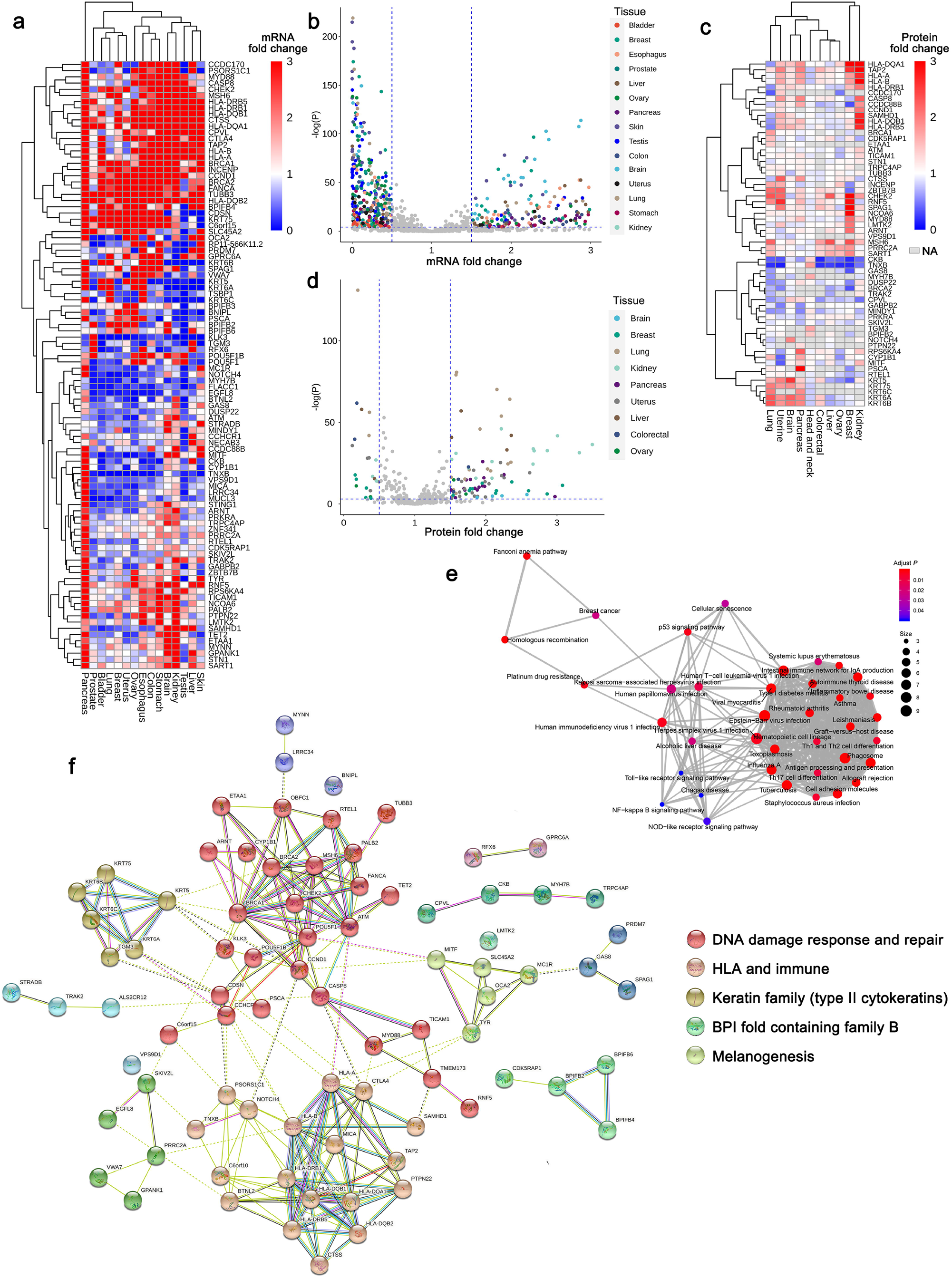
(a) Heatmap of fold change (FC) values to compare gene expression between TCGA tumor tissues and GTEx healthy normal tissues. (b) Volcano plot for the FC values and -log(P) values for comparison of gene expression. (c) Heatmap of FC values to compare protein abundance between CPTAC tumor tissues and adjacent normal tissues. NA: not available. (d) Volcano plot for the FC values and -log(P) values for comparison of protein abundance. (e) KEGG pathway network from the enrichment analysis of the pleiotropic genes. (f) Protein-protein interaction network of the signal genes and pleiotropic genes.

Further, we performed a KEGG pathway enrichment analysis for the identified genes. Numerous immune-related pathways were identified, such as Th1 and Th2 cell differentiation (*P* = 1.46 ×10^−3^), Th17 cell differentiation (*P* = 2.63 ×10^−3^), and inflammatory bowel disease (*P* = 3.94×10^−4^), as well as the classical cancer-related pathways, including cell adhesion molecules (*P* = 1.29 × 10^−5^), platinum drug resistance (*P* = 6.14×10^−4^), p53 signaling pathway (*P* = 6.14×10^−4^), and NF-kappa B signaling pathway (*P* = 0.018) (Figure 3e, Table S9).

Proteomic data were collected across ten tissue types from The National Cancer Institute’s Clinical Proteomic Tumor Analysis Consortium (CPTAC). We observed moderate differences by comparing protein abundance in tumor and adjacent normal tissues. Interestingly, some proteins showed similar patterns with the corresponding gene expression, such as *HLA* family genes and *CHEK2*. However, a reverse trend was also found for some genes. For example, *BRCA2* was down-regulated in proteomics of tumor tissues (Figure 3c, 3d).

Using the STRING database to integrate all known and predicted proteins-protein interactions, we identified two large clusters: one was related to the HLA (e.g., HLA family genes, *MICA, BTNL2*) and immune function (e.g., *CTLA4, CTSS, SAMHD1*); another was related to DNA damage response and repair (e.g., *BRCA2, ATM, CHEK2, MSH6*) (Figure 3f). In addition, three small clusters were also identified, including the keratin family, BPI fold containing family B, and melanogenesis.

### Identify high-risk population based on rare variants

We developed exome-wide risk scores (ERS) to identify high-risk population based on rare variants with minor allele frequency (MAF) < 0.05 for incident cancers. The discovery set (WES-300k population) was used for risk score training, while the replication set (WES-150k) was used for external validation. After screening using the least absolute shrinkage and selection operator (LASSO) with 10-fold cross-validation, we included a moderate number of rare variants in ERS (median [interquartile range (IQR)]: 103 (82-199)), ranging from 39 variants (thyroid) to 585 variants (skin) (Table S10).

We performed a risk stratification analysis to identify the high-risk populations susceptible to specific cancer types. We defined three risk levels: extremely high-risk (top 5% ERS), high-risk (5%-25% ERS), and low-risk (bottom 75% ERS), which might be applicable to different medical screening strategies. The ERS could stratify the cancer absolute incidence risk significantly in the replication set (Figure 4) and the whole UKB-450k population (Figure S4) (all log-rank *P*<2.2×10^−16^). Using the low-risk population as the reference, the extremely high-risk persons had hazard ratios (HRs) ranging from 4.52 to 9.92 [median (IQR): 7.20 (6.44-7.85)], which outperformed PRS [HR median (IQR): 2.44 (1.95-2.80)]. The high-risk persons had HRs ranging from 1.37 to 4.43 [median (IQR): 1.97 (1.75-2.33)), while the PRS had HRs ranging from 1.11 to 2.15 with median (IQR): 1.56 (1.41-1.79) (Figure 5a, 5b). Thus, the ERS had satisfactory performance in cancer risk stratification, especially for the extremely high-risk persons that were frequently risk allele carriers. The distributions of ERS in each cancer type were shown in Figure S5. Most people did not carry the causal alleles, while only a few were high-risk carriers. The density plot of cases and controls indicated the cancer cases had obvious larger ERS than controls (Figure S6).

**Figure 4.**
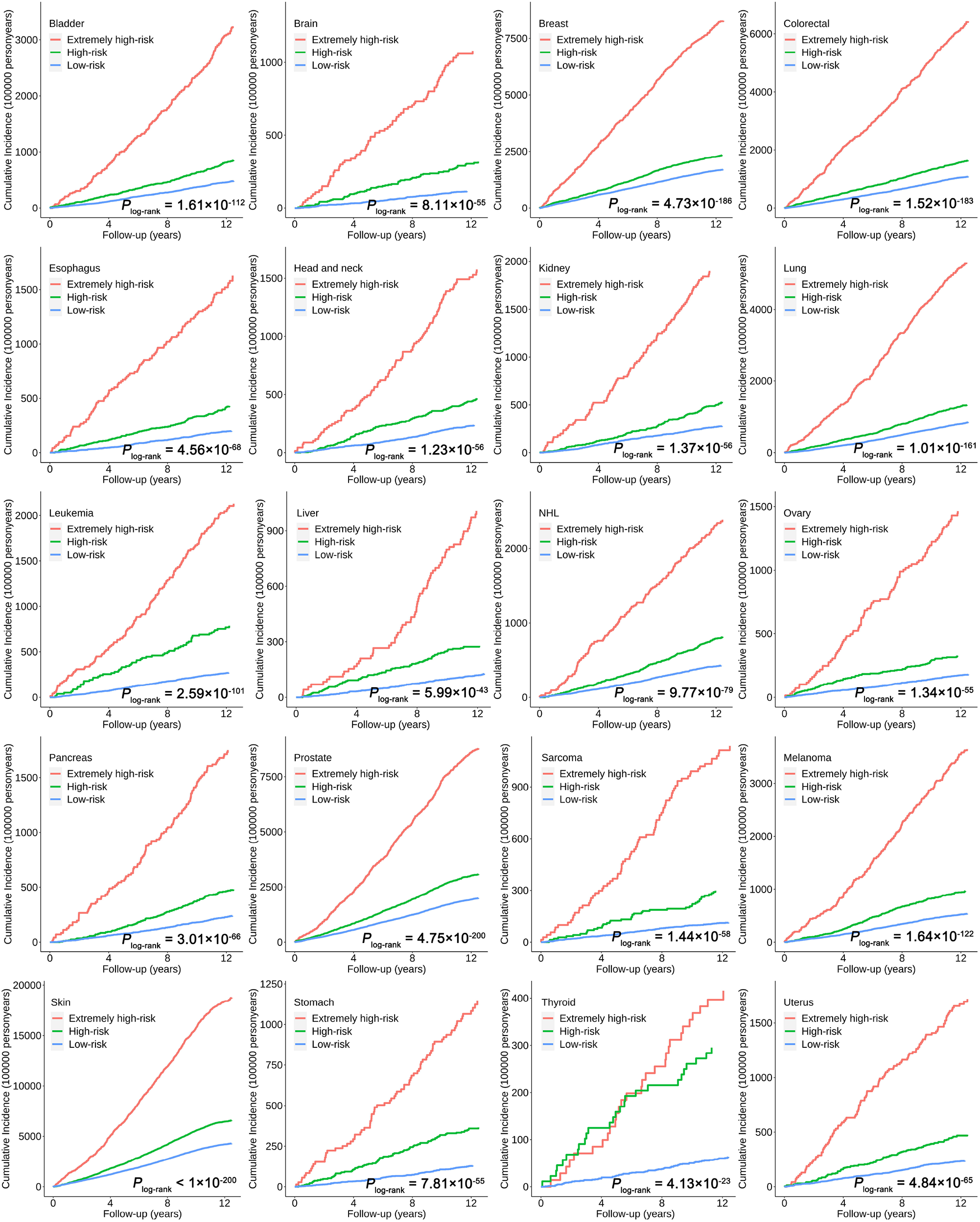
Cumulative cancer incidence plot for the 20 cancer types in the replication set (WES-150k). The red line indicates the extremely high-risk persons, the green line indicates the high-risk population, and the blue line indicates the low-risk persons. The *P* values were calculated using the log-rank tests.

**Figure 5.**
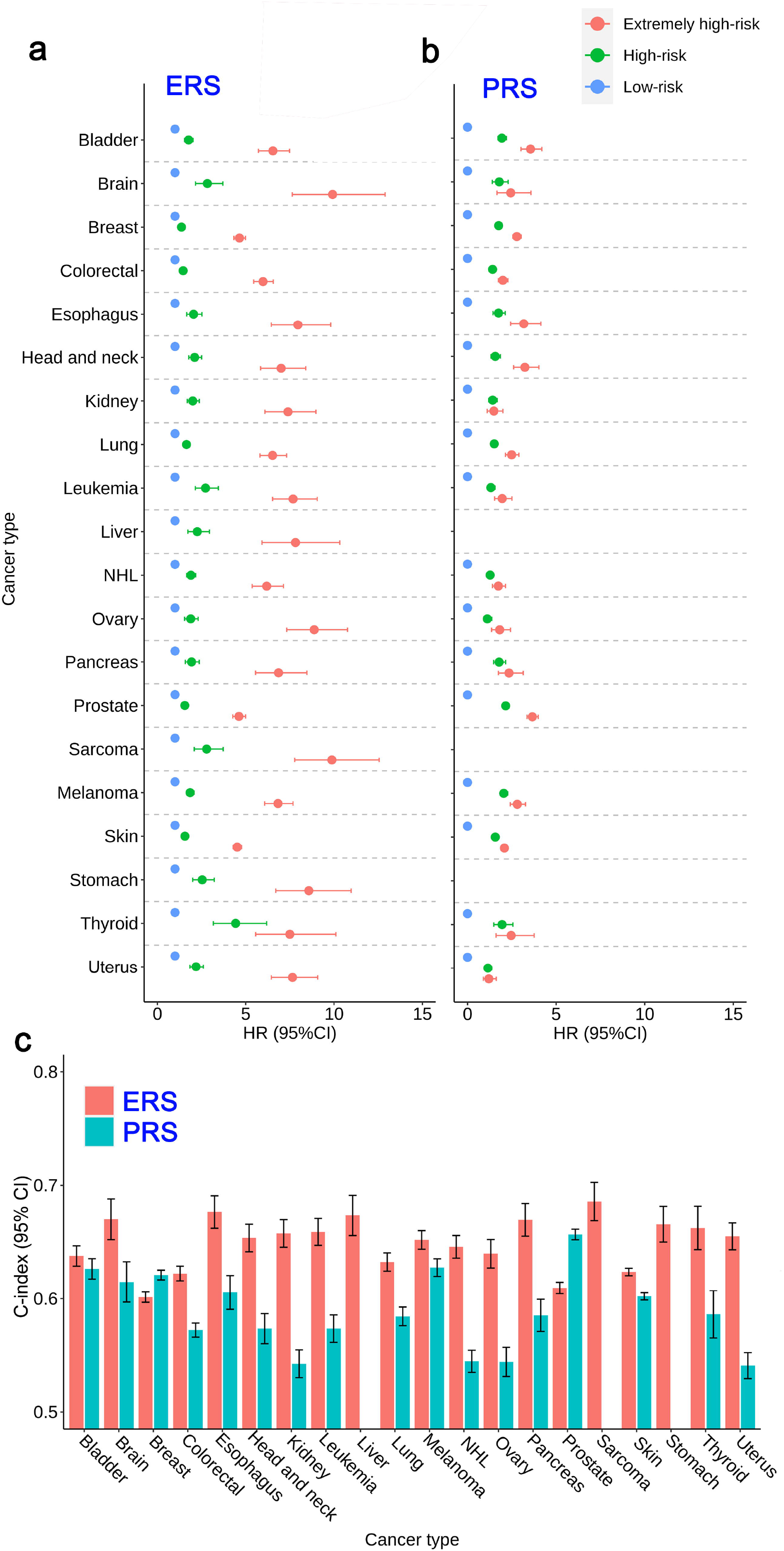
(a) The dot plot of the hazard ratios (HRs) and 95% confidence intervals (CIs) of exome-wide risk scores (ERS). The low-risk subgroup (blue dot) was set as the reference group. The red dot indicates the extremely high-risk persons, the green dot indicates the high-risk population. (b) The dot plot of the HRs and 95% CIs of polygenic risk scores (PRS) for 17 cancer types, while liver, stomach, and sarcoma cancers did not have reported PRSs. (c) The C-index values of ERS and PRS generated by the Cox regression model.

Further, we evaluated the discrimination abilities of ERS and PRS using the C-index for ten-year cancer incidence. The C-index values in the replication set were stable, ranging from 0.601 to 0.686 [median (IQR): 0.655 (0.636-0.667)], outperforming the traditional PRS [median (IQR): 0.585 (0.572-0.614)] (Figure 5c). Through Spearman correlation analysis, we found low correlation between ERS and PRS (average *r*_*s*_ = 0.016), indicating the complementary predictive value for the rare exome variants (Table S11).

## Methods

### Study population and phenotype definition

The UK Biobank (UKB) is a population-based prospective cohort of individuals aged 40–69 years, enrolled between 2006 and 2010. The work described herein was approved by the UK Biobank under application no. 83445. All the phenotype data were accessed in March 2022.

Health-related outcomes were ascertained via individual record linkage to national cancer and mortality registries and hospital in-patient encounters. Cancer diagnoses were coded by International Classification of Diseases version 10 (ICD-10) codes. Individuals with at least one recorded incident diagnosis of a borderline, in situ, or primary malignant cancer were defined as cases collected from data fields 41270 (Diagnoses - ICD10), 41202 (Diagnoses - main ICD10), 40006 (Type of cancer: ICD10), and 40001 (primary cause of death: ICD10). Finally, we analyzed 20 primary cancer types with overall cases > 500, including bladder, brain, breast, colorectal, esophagus, head and neck, kidney, lung, leukemia, liver, non-Hodgkin’s lymphoma (NHL), ovary, pancreas, prostate, sarcoma, melanoma, skin (non-melanoma), stomach, thyroid, uterus cancers.

To ensure the robustness of the results and validate the risk stratification model, we used the WES-300k population released on Sep 28^th^ 2021 as the discovery set and the remaining WES-150k population additionally released on Oct 29^th^ 2021 as the replication set. We included only participants of European ancestry. The demographic and cancer characteristics were described in Table S1. All the data analyses were performed on DNAnexus Research Analysis Platform (RAP).

### Quality control for the genetic variants

Whole-exome sequencing data for UKB participants were generated using the IDT xGen v1 capture kit on the NovaSeq6000 platform. The UK Biobank WES 450k release includes CRAM and gVCF files processed using the OQFE protocol ^19^. Single-sample variants were called from OQFE CRAMs with DeepVariant 0.0.10 employing a retrained model and are provided as single-sample gVCFs. All gVCFs were aggregated with GLnexus 1.2.6 using the default joint-genotyping parameters for DeepVariant. The OQFE protocol maps to a full GRCh38 reference version including all alternative contigs in an alt-aware manner. Genotype depth filters (SNV DP≥7, indel DP≥10) were applied prior to variant site filters requiring at least one variant genotype passing an allele balance filter (heterozygous SNV AB>0.15, heterozygous indel<0.20). The detailed parameters were described in Category 170 of the UKB showcase. In addition, we filtered out the variants with low allele counts if a variant had minor allele count (MAC) < 3 in the discovery set or MAC < 2 in the replication set.

### Exome-wide association for single variants and gene levels

Single-variant and gene-based association analyses were performed using SAIGE v1.0.5 ^20,21^. SAIGE is a toolkit developed for genome-wide association tests in biobank level datasets, which uses saddlepoint approximation to handle extremely case-control imbalance of binary traits and linear mixed models to account for sample relatedness. The variant-level association tests included high-quality and reliable variants with MAC≥10. The exact inclusion criteria for single variants were: (i) MAC ≥3 in both cases and controls in the discovery set; (ii) MAC≥2 in both cases and controls in the replication set. The variants were functionally annotated using Variant Effect Predictor (VEP) software ^22^. In addition to the coding variants that alter protein sequences, synonymous variants could also disturb the level of mRNA expression and have non-neutral functions ^23,24^. Thus, variants annotated as putative loss-of-function (LoF, including nonsense, splice site, and frameshift variants), missense, and synonymous were included in the analysis. Independent variants were pruned out using the PLINK v1.9 clump function (*--clump-r2 0*.*50 --clump-kb 500*).

For gene-based analysis, we included rare and ultra-rare variants with minor allele frequency (MAF) < 0.05, 0.01, or 0.001. Three genetic models were considered: LoF, LoF+missense, LoF+missense+synonymous. Of all the combinations, we reported the association results with the lowest *P* value to collectively capture a wide range of genetic architectures ^25^. The effect sizes and 95% confidence interval (CI) of genes were estimated by burden tests.

In all the association analyses, we adjusted the covariates including age, gender (excluding sex-specific tumors), Body Mass Index (BMI), smoking status (binary), drinking status (binary), and the top 10 principal components (PCs). Meta-analysis was used to summarized the results between discovery and replication sets for single variants by METAL software ^26^. At the same time, the gene-based *P* values were aggregated by aggregated Cauchy association test (ACAT) method ^27^.

We reported the significant associations when the variants/genes meet the following criteria: (i) *P* < 0.05 in both discovery and replication sets; (ii) reach genome-wide significant level (*P* < 5×10^−8^) in the meta-analysis for variant-level or pass Bonferroni correction threshold *P* < 2.5×10^−6^ (nearly 20,000 protein-coding genes tested) for gene-level.

### Shared genetics analyses to identify potential genes across cancers

To discover more potential variants associated with multiple cancers, we applied a cross-trait meta-analysis using Association analysis based on SubSETs (ASSET) ^28^. ASSET is a statistical tool specifically designed to be powerful for pooling association signals across multiple cancers when true effects may exist only in a subset of the cancers. We considered both one-sided (one-directional) and two-sided (bidirectional) ASSET. Variants with association *P* < 1×10^−4^ in any cancer type were included. Variants were considered significant while reaching *P*_overall_ < 5×10^−8^ and *P* < 0.05 for each side in bidirectional tests.

### Comparison analyses for gene expression in tumor and healthy normal tissues

The UCSC Toil Recompute Compendium provides processed transcript-level RNA-Seq data from TCGA tumor tissues and GTEx healthy normal tissues quantified using a unified computational pipeline to remove computational batch effects. We used this data to perform comparative analysis across tumor and health normal tissues from both projects ^29^. All the gene expression values were normalized to transcripts per million (TPM) and then logarithmically transformed. After filtering, 15 tissue types with sample size > 100, including 7,085 TCGA tumor tissues and 4,311 GTEx normal tissues, were analyzed (Table S12). We used Student’s t-test (*P*_bonferroni_<0.05) and fold change (FC>1.5 or FC<0.5) to identify the differential gene expression between tumor and healthy normal tissues.

### Pathway enrichment analysis

We collected the pathway information with gene sets from the KEGG database, containing a total of 186 pathways up to March 2022. All enrichment analyses were performed using the R package *clusterProfiler* ^30^.

### Comparison analyses for protein abundance in tumor and adjacent normal tissues

The National Cancer Institute’s Clinical Proteomic Tumor Analysis Consortium (CPTAC) is a national effort to accelerate the understanding of the molecular basis of cancer through the application of large-scale proteome and genome analysis ^31^. Eleven cancer types with available protein abundance data of 1,270 tumor tissues and 845 adjacent normal tissues were collected from ten tissue types (Table S13). The differential proteins were identified following the same criteria with gene expression.

### Protein-protein interaction analysis

To further understand the protein-protein interactions, we used the STRING database, which considered both physical interactions as well as functional associations ^32^. The protein interaction network was clustered into different colors using Markov Clustering (MCL).

### Polygenic risk score generation

The polygenic risk score (PRS) aggregates the effects of numerous genetic variants into a single number which predicts genetic predisposition for a phenotype. To investigate the association of PRS with cancer risk, we generated the PRS based on previous literature reports. The SNP information and score weights were collected from the PGS Catalog database ^33^. PGS Catalog is an open database of published PRSs, covering >2,000 scores for various phenotypes. If multiple PRSs were reported for one cancer, we selected the one generated from the largest sample size. After filtering, 17 PRSs were collected and generated in UKB imputed genotype population (data field 22828: Imputation from genotype), except for liver, stomach, and sarcoma cancers that did not have reported PRSs (Table S14).

### Development and validation of the risk stratification models based on rare variants

We developed an exome-wide risk score (ERS) for population risk stratification based on rare variants (MAF<0.05) for each cancer separately. The analyses were restricted to incident cancers. To perform independent training and validation phases, the selected variants and weights were determined using the association information in the discovery set.

We select the rare genetic variants reaching *P*<1×10^−4^ in the discovery set as the first step. The variants identified in this analysis were further screened through penalized regression using the least absolute shrinkage and selection operator (LASSO) after 10-fold cross-validation to reduce the overfitting and collinearity problem. When high correlation exists, variants representing independent loci with the strongest statistical significance were retained. The ERS was generated as: 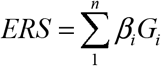, where *β*_*i*_ denoted the coefficient of the i^th^ variant *G*_*i*_ calculated by SAIGE in the discovery set.

In the validation phase, the variant panel with previously determined weights was used to generate ERS in the replication set.

We used person-year to describe the absolute cancer incidence risk, which was defined as the time gap from the date of cohort enrollment to cancer diagnosis or the last follow-up, whichever came first. Hazard ratios (HRs) and 95% confidence interval (CI) were used to evaluate the association between ERS and cancer risk based on Cox proportional hazards models, adjusting for age, sex (excluding sex-specific cancer), and top ten principal components. We compared effect sizes of ERS for cancer risk based on the top 5% (extremely high-risk), 5∼25% (high-risk), and bottom 75% (low-risk) percentile of ERS. The discrimination performance of the risk scores were evaluated by Harrell’s C-index.

## Discussion

In this study, we comprehensively evaluated the susceptibility between genetic variants on human exome and 20 primary cancer types in approximately 420,000 UKB participants of European ancestry. To our knowledge, this is the first exome-wide pan-cancer study including almost the whole UKB population, which could improve the statistical power compared with some previous studies using the early-phase UKB 200k population ^34,35^. Trans-omics analyses were performed to evaluate the functional evidence of identified signals, including genomics, transcriptomics, and proteomics. Moreover, through establishing the independent discovery and replication sets, the signals identified and the risk stratification models could be validated externally to ensure their robustness, especially for the rare variants with smaller MAF.

Our first major finding discovers exome-wide signals associated with multiple cancers. In the ExWAS analyses, the identified protein-coding variants in *CHEK2* (known as c.1100del), *BRCA1/2, ATM* have been reported associated with cancers in previous studies ^36^. Based on such a large-scale population, novel variants and genes were also identified. For example, the missense variant rs6998061 in *POU5F1B*, which was predicted as a possibly damaging SNV by VEP, was associated with prostate and colorectal cancers. *POU5F1B* was a protein-coding gene highly homologous to *OCT4* ^37^, which was recently shown to be transcribed in cancer cells. It has been found related to tumorigenicity and tumor growth *in vivo* and could promote angiogenesis and cell proliferation and inhibits apoptosis in cancer cells ^38^. The stop lost variant rs387907272, predicted as probably damaging in *MYD88*, was associated with leukemia and NHL. *MYD88* encodes a cytosolic adapter protein that plays a central role in the innate and adaptive immune response ^39^. It functions as an essential signal transducer in the interleukin-1 and Toll-like receptor signaling pathways ^40^. Moreover, mutations in *MYD88* could activate NF-κB and its associated signaling pathways, thereby promoting B-cell proliferation and survival ^41^. Thus, this germline mutation might be a promising target for clinical implications. Moreover, we identified multiple SNVs located in 16q24.3 that were shared between skin cancer and melanoma, including *VPS9D1, MC1R*, and *TUBB3. VPS9D1*, a protein-coding gene that affects protein binding activity, was significantly associated with skin cancer and melanoma. Although its role in cancer has not been reported, its antisense *VPS9D1-AS1* could promote tumorigenesis and progression by mediating micro RNAs via the Wnt/β-catenin signaling pathway in multiple cancers ^42,43^. Human *MC1R* has an inefficient poly(A) site allowing intergenic splicing with its downstream neighbor *TUBB3*, which were involved in melanogenesis. Melanogenesis is a key parameter of differentiation in melanocytes and melanoma cells that could affect the treatment of pigmentary disorders ^44^. Therefore, the SNVs and genes we identified had remarkable biological functions that were practical for precise clinical implications.

Our second major finding identified the pleiotropic variants shared among cancers. We observed strong functional evidence for the identified genes from KEGG pathway network and protein-protein interaction network through trans-omics analyses for gene expression and protein abundance. We found several essential function modules from proteomics. The DNA damage response and repair agents are widely used in clinical oncology given the expanding role of immune checkpoint blockade as a therapeutic strategy ^45^. The HLA locus, located on chromosome 6, is among the most polymorphic regions of the human genome ^46^. HLA dysfunction is deeply involved in the immune evasion events in the development and progression of certain cancers ^47^. A previous study has reported that somatic mutation in HLA was associated with multiple cancers ^48^; we hereby demonstrated that germline mutation in HLA was also relevant. In addition, the keratin family and BPI fold containing family B (BPIFB) were also related to cancers that had certain biological functions ^49,50^.

Our third major finding improves the ability for high-risk population identification. It is widely recognized that early screening for cancers is most likely beneficial when the target tumor type has relatively uniform biology and a slower rate of progression ^2^. Targeting on high-risk populations with appropriate strategies for early detection could get remarkable benefits of mortality reduction ^51-53^. However, the selection of individual to be screened is a complex procedure, with difficulty accurately identifying high-risk persons who are most likely to benefit from screening. Because cancer is heritable, PRS is emerging as the quantitative measurement for individual genetic risk. However, the heritability for common variants identified in GWAS is limited, while the contribution of rare variants could not be ignored. Therefore, we leveraged the rare variants to construct the ERS to offset this limitation. By evaluating C-index and risk stratification, we demonstrated the added values of rare variants.

ERS could be combined with specific tumor screening strategies, suggesting that people with extremely high risk should be screened frequently (e.g., once a year), and those with high risk should be screened regularly (e.g., once every three years), which is expected to further reduce the cancer mortality. Therefore, ERS is expected to serve as an informative benchmark to incorporate the PRS and baseline information that have been used in cancer risk assessment.

Our work has several strengths. First, we comprehensively evaluated the exome-wide genetic variants in 20 cancer types on variant and gene levels among 420k participants and analyzed the cross-cancer pleiotropy through cross-trait meta-analysis. Second, we explored the relationship between identified genes and cancers at multi-omics levels, including genomics, transcriptomics, and proteomics. The trans-omics analyses revealed that the identified signals were functional. Third, we focused on the high-risk population identification based on exome-wide variants, while few studies developed risk scores using rare variants. We demonstrated the stable performance of ERS across pan-cancer in the replication set, especially for its ability to identify the extremely high-risk persons. Therefore, the ERS might serve as a complementary genetic risk assessment tool combined with the existing screening guidelines.

It is essential to acknowledge the limitations of our study. First, this study was conducted in the UKB population only. Although we established a discovery set and replication set, it is not strictly independent validation. Therefore, future large-scale population studies should be conducted to replicate these findings. Second, we focused on individuals of European ancestry only. Moreover, it is essential to evaluate the associations of variants and performance of ERS in non-European populations. Third, we mainly investigated the genetic effects of population risk stratification. However, the contribution of environmental factors should not be ignored. Well-established risk models incorporated with environmental factors, PRS, and ERS should be developed for specific cancer.

In conclusion, our study provides novel insights into human exomes and rare variants through comprehensive analyses of genetic susceptibility to human cancers and subsequent target analyses on specific genes and risk stratification.

## Data Availability

UK Biobank data is available from https://www.ukbiobank.ac.uk/. TCGA data is available from https://portal.gdc.cancer.gov/. GTEx data is available from https://www.gtexportal.org/home/. CPTAC data is available from https://pdc.esacinc.com/pdc/pdc.

## Code Availability

The R software codes that support our findings are available from the corresponding author by a reasonable request.

## Supplementary Figures

Figure S1. Q-Q plots of single variant tests for each cancer

Figure S2. Q-Q plot combining all the P values of pan-cancer

Figure S3. Manhattan plot for the single variant analyses of each cancer

Figure S4. Cumulative cancer incidence plot for the 20 cancer types in the whole UKB-450k population.

Figure S5. Histogram of the exome-wide risk scores (ERS) in each cancer

Figure S6. Density plot of the exome-wide risk scores (ERS) in cases and controls

## Supplementary Tables

Table S1. Demographic characteristics in the UK Biobank cohort

Table S2. Association results for independent single variants with P<5×10^−8^ in the whole UKB-450k population

Table S3. Association results for single variants with P<5×10^−8^ in at least two cancer types

Table S4. Association results for genes with P<2.5×10^−6^ in the whole UKB-450k population

Table S5. Association results for genes with P<2.5×10^−6^ in at least two cancer types

Table S6. Cross-trait meta-analysis results for single variants with one-directional effects

Table S7. Cross-trait meta-analysis results for single variants with bidirectional effects

Table S8. Number of independent shared genetic variants in the cross-trait meta-analysis

Table S9. KEGG enrichment analysis for the genes identified in cross-trait meta-analysis

Table S10. Model parameters for the exome-wide risk scores (ERS) in 20 cancer types

Table S11. Correlation analysis for ERS and PRS

Table S12. Number of tissues included in the comparison analyses for gene expression in tumor and healthy normal tissues

Table S13. Number of tissues included in the comparison analyses for protein abundance in CPTAC

Table S14. Information of the selected polygenic risk score (PRS) in PGS catalog

