## Supplemental figures for "Exome-wide association studies discover germline mutation patterns and identify high-risk populations in human cancers"

**Supplementary Figures**

### Figure S1. Q-Q plots of single variant tests for each cancer

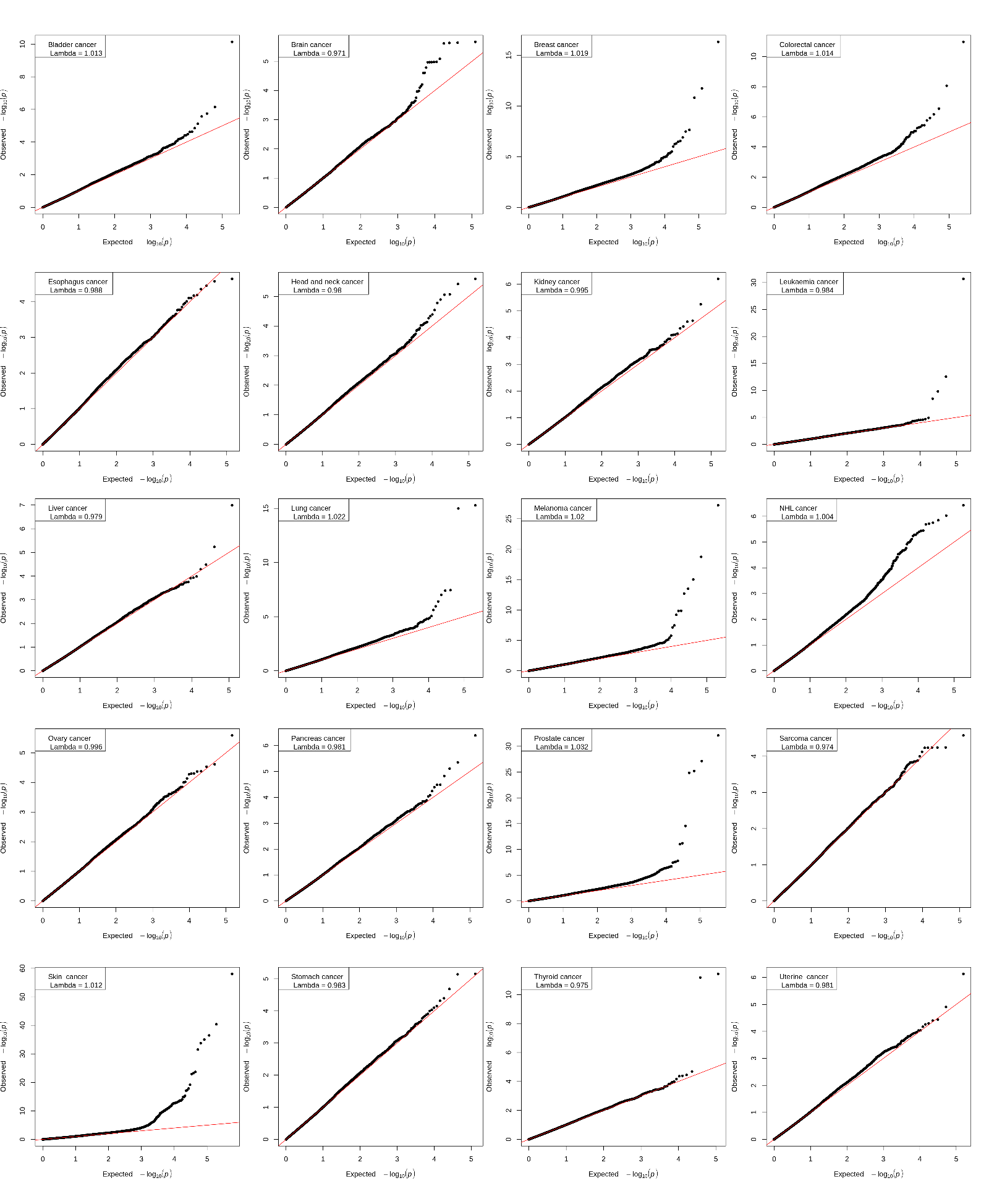

### Figure S2. Q-Q plot combining all the *P* values of pan-cancer

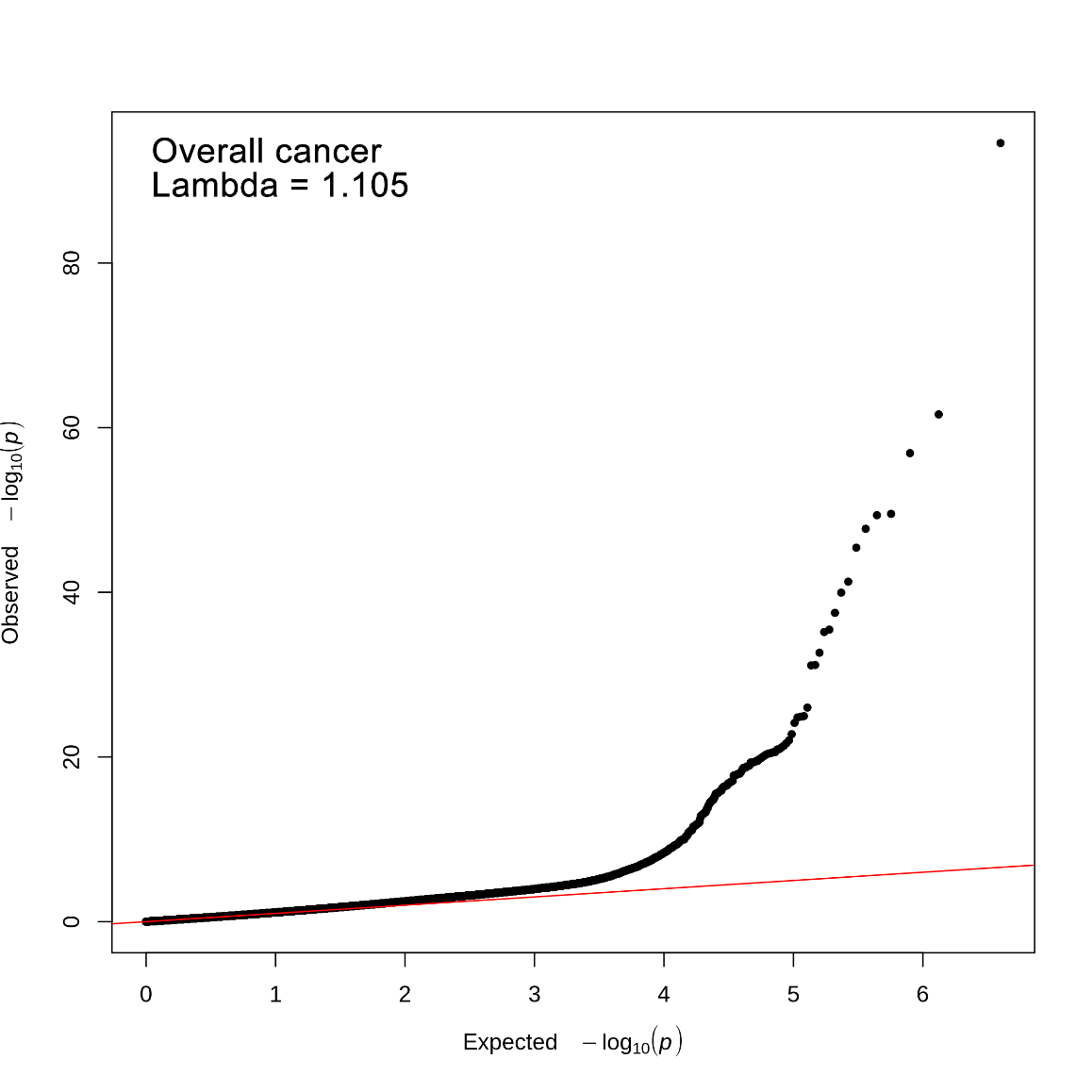

### Figure S3. Manhattan plot for the single variant analyses of each cancer

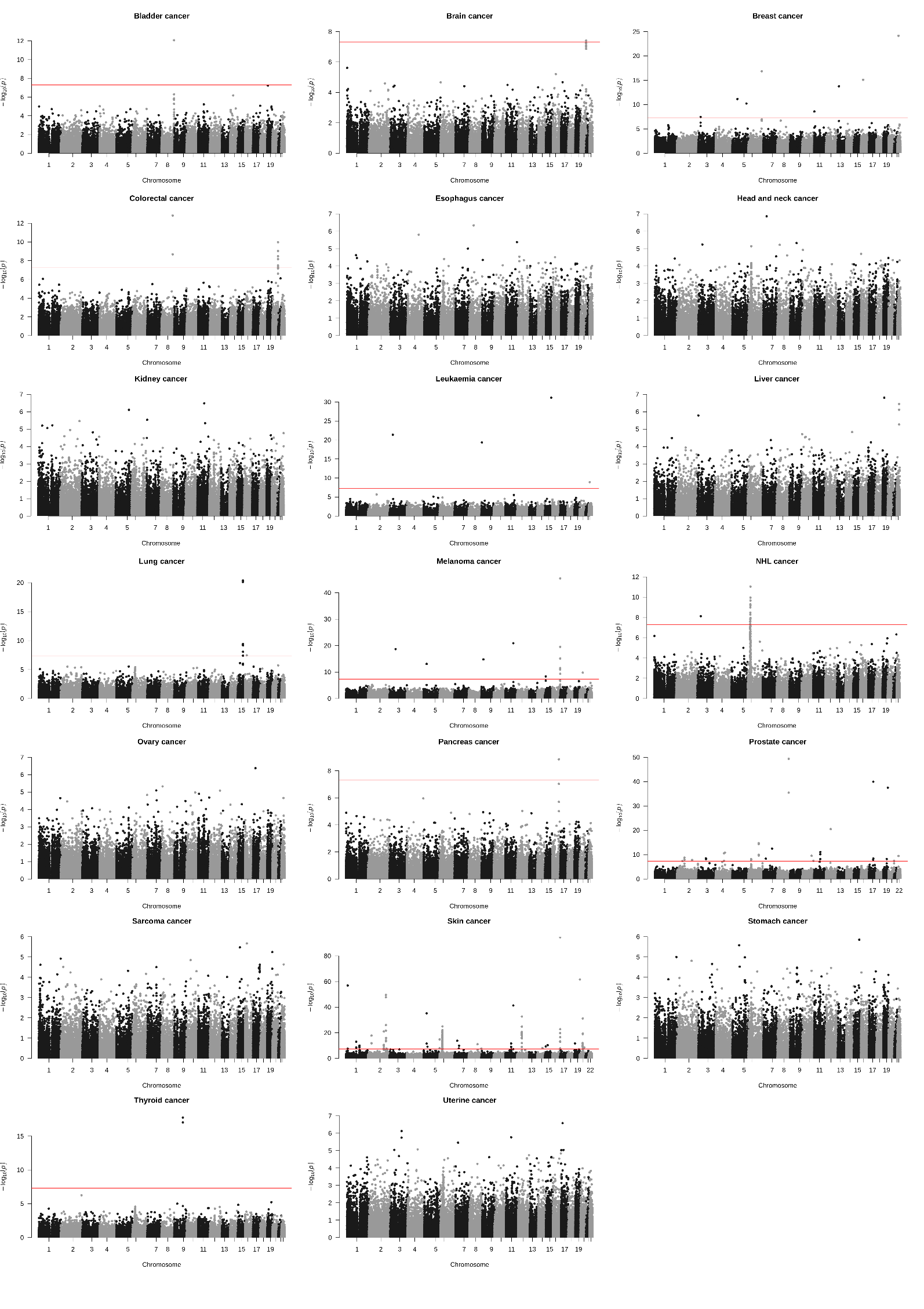

### Figure S4. Cumulative cancer incidence plot for the 20 cancer types in the whole UKB-450k population

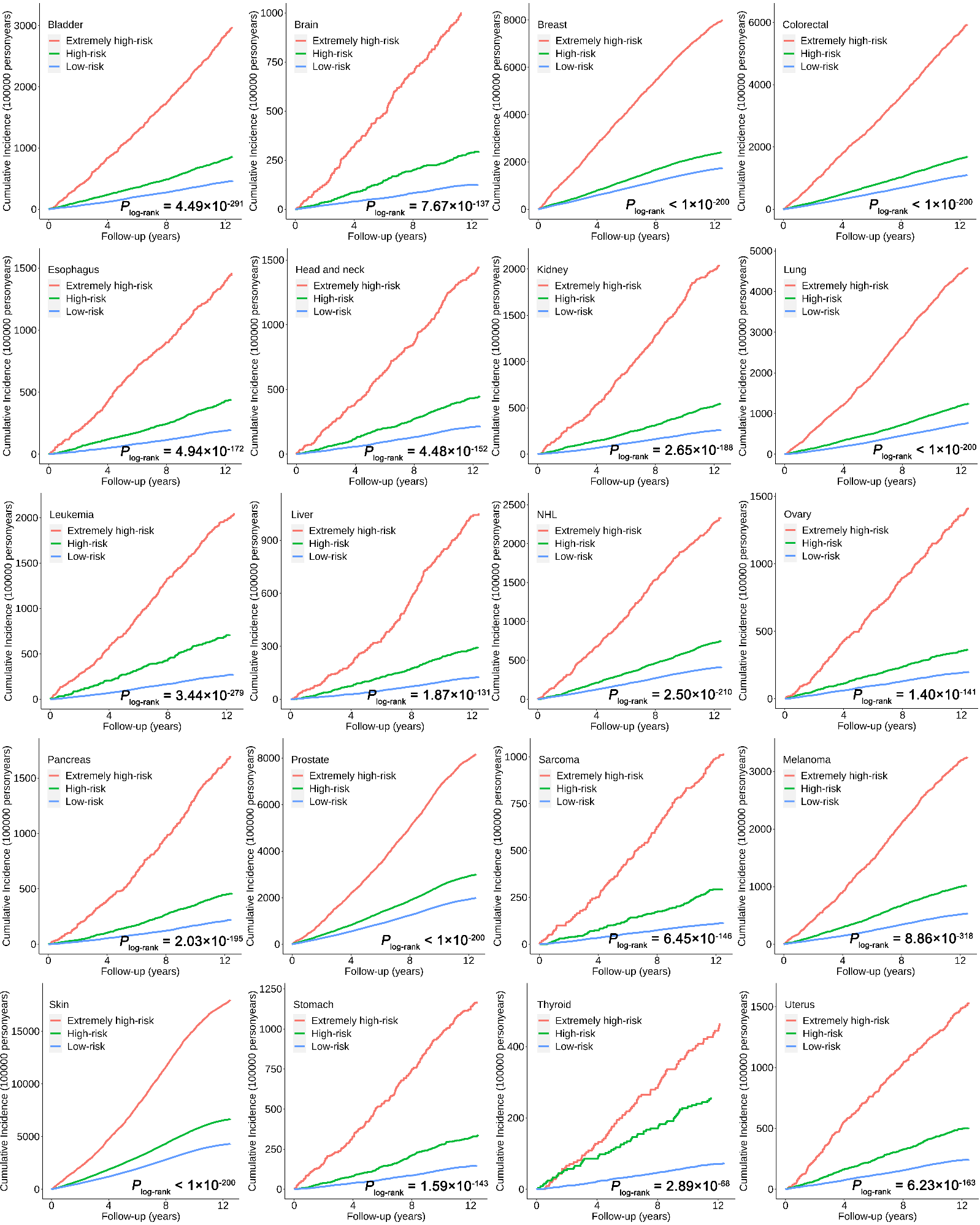

### Figure S5. Histogram of the exome-wide risk scores (ERS) in each cancer

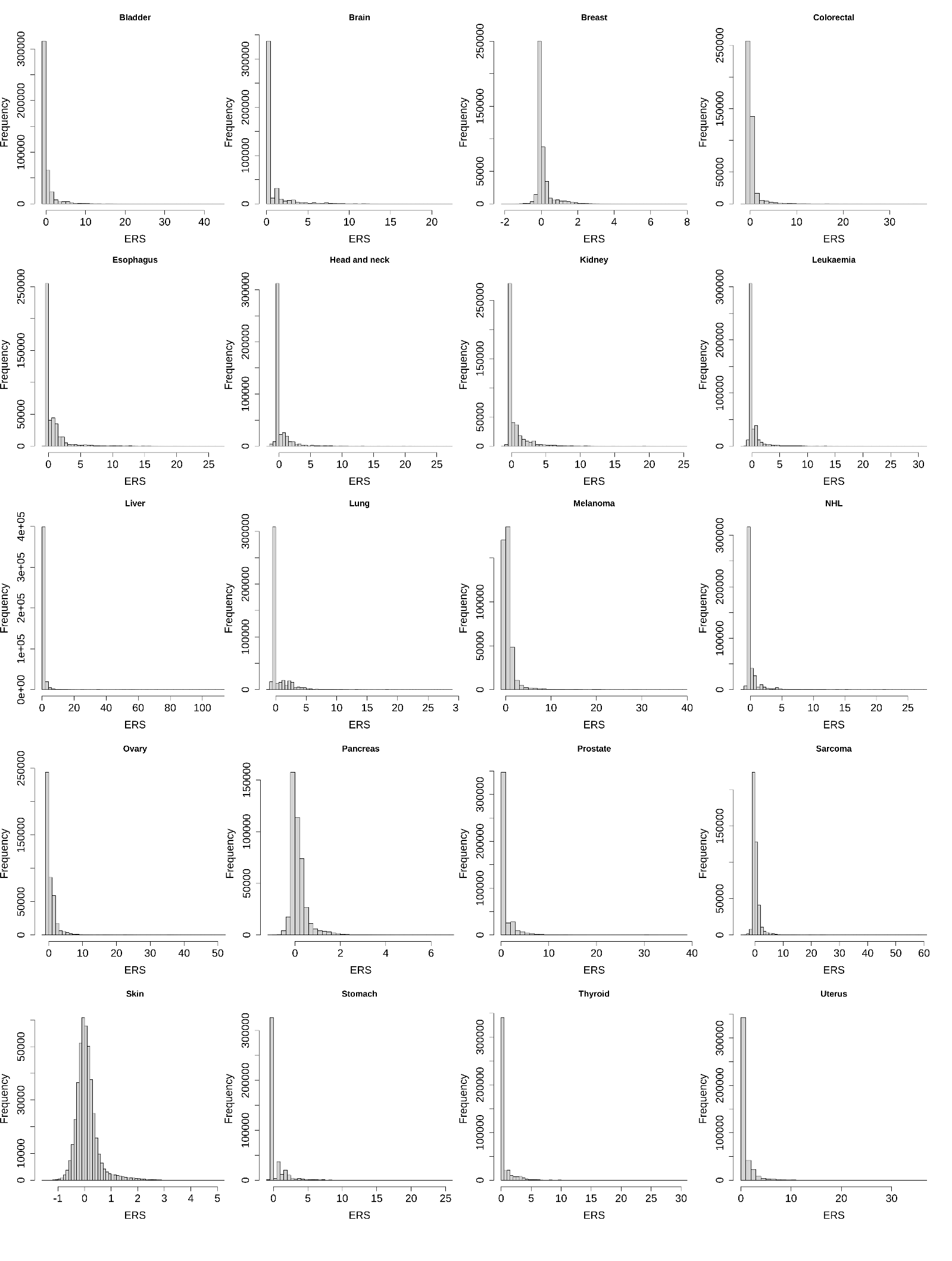

### Figure S6. Density plot of the exome-wide risk scores (ERS) in cases and controls

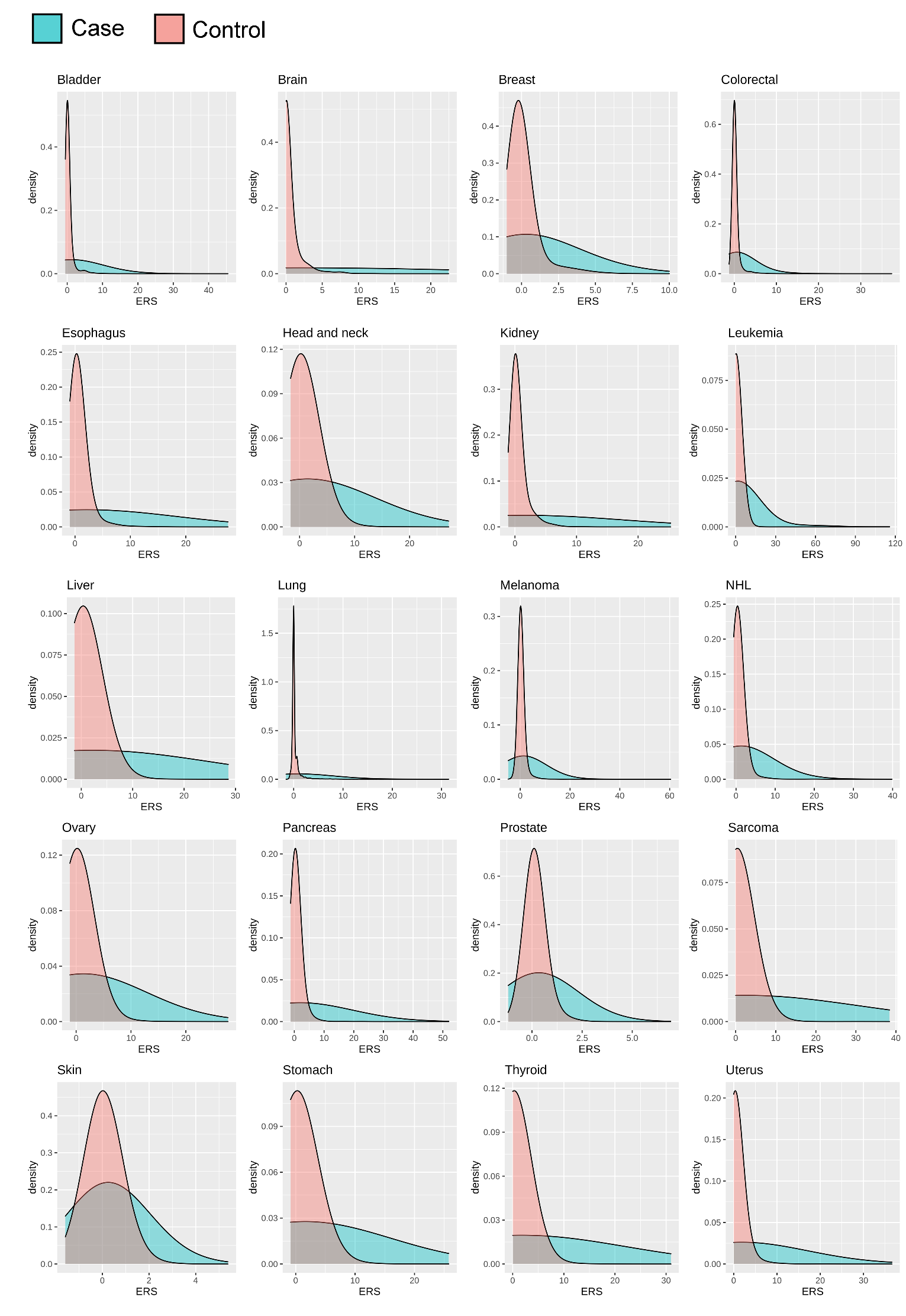
